# Investigation research on the demand of pre-job training for nursing students in higher vocational colleges

**DOI:** 10.1101/2021.07.10.21260295

**Authors:** Li Liu, Xiaojing Qiu, Yahui Li, Junzheng Yang

## Abstract

**Objectives:** To investigate the pre-job training demand of nursing students in higher vocational colleges, lay a foundation for the development of scientific and standardized pre-job training program and help nursing students smoothly transition to clinical practice state from student status.

**Methods:** A self-designed questionnaire was used to investigate 822 nursing students of grade 2018 in our school from four aspects: the necessity of pre-job training, the demand content of pre-job training, the demand of skill enhancement program, and the stress source of pre-internship.

**Results:** Demand rate of the pre-job training of nursing students in the college was 99.5%, and main demand of the training content were nursing skill training (96.8%), nursing safety and occupational protection (95.1%), nursing theory training (92.9%); the main demand of nursing skills enhancement program were closed intravenous infusion (90.3%), (intradermal, subcutaneous, intramuscular) injection (87.8%), cardiopulmonary resuscitation (81.8%), and indwelling catheterization (77.5%); the main stress sources of pre-internship was lack of confidence in theory and skills, accounts for 61.1% in total 760 students.

**Conclusion:** The pre-job training courses in higher vocational colleges should be close to the clinical needs of students, it is necessary to carry out pre-job training courses according to main demand of the training content, the main demand of nursing skills enhancement program and the main stress sources of pre-internship in the college; this article may provide reference for our school and other vocational colleges to develop pre-job training program.

## 1. Introduction

As an important training base of nursing professional skilled talents, the teaching staff of higher vocational colleges must scientifically formulate talent training programs, execute high standard and strict requirement for students in nursing teaching and nursing training, cultivate more excellent nursing talents for the society. Pre-job training is an important course for higher vocational nursing students before they enter the hospital for clinic practice. Targeted standardized pre-job training for nursing students can make them better transform their roles and adapt to the new work, so as to improve their professional quality and provide better service for patients^[1]^.

At present, the domestic pre-job training of nursing mainly includes pre-job training in schools and hospitals. The training contents include basic theoretical knowledge training, common clinical nursing operation technology training, professional theory and practice ability training. Since the establishment of nursing specialty in 2015, our school has been taking pre-job training as a course throughout the whole second semester of sophomore year. The training effect has been affirmed by students and practice hospitals, and the satisfaction rate is 99.3%, but there are still some shortcomings. In order to better improve the pre-job training course in our college, we conducted semi-structured interviews with the interns who had finished their internships in our school and preliminarily understand the advantages and disadvantages of our nursing students in the hospital internship, combined with the feedback from the practice hospital, we developed an electronic questionnaire on the pre-job training needs of nursing students, and conducted a questionnaire survey on the pre-job training needs of nursing students through the network teaching platform in our school, aim to develop a more scientific and standardized pre-job training program, make the pre-job training content close to the hospital practice requirements, meet the physical and mental needs of students and help nursing students to complete the role transformation to enter clinical practice.

## 2 Investigation objects and Methods

### 2.1 Investigation objects

822 nursing students of grade 2018 were investigated by online questionnaire through the online teaching platform of our university in 2020. A total of 760 valid questionnaires were collected, and the effective rate was 92.45%. The average age of the investigation objects was 20.17±0.89 years old, there were 98 male students, accounting for 12.9%, and 662 female students, accounting for 87.1%.

### 2.2 Investigation methods

1. Firstly we set up the 2018 pre-job training needs investigation group through the network teaching platform of our school, totally 822 nursing students were investigated in our school. Students can complete the questionnaire in the mobile terminal learning software.
2. In this study, a self-designed questionnaire was used to investigate the pre-job training needs of nursing students. The questionnaire was designed by the researchers of the research group through literature review, student interviews, expert consultation, combined with the training program for new nurses in 2016 and the actual situation of pre-job training in our school. It includes the necessity of pre-job training, demand content of pre-job training, the demand of skill enhancement program and stress source of pre-internship.

## 3. Statistical analysis

In this study, SPSS 23 Statistical software was used and multiple response analysis was used to analyze multiple choice questions.

## 4. Results

### 4.1 Investigation results of the necessity of pre-job training courses

Firstly, we investigated the students’ opinion on the necessity of pre-job training. There were 583 students thinking that pre-job training was very necessary, accounts for 76.7% of the total number of students surveyed; there were 173 students thinking that pre-job training was necessary, accounts for 22.8% of the total number of students surveyed; and 0.5% students thought that pre-job training was not necessary, the result was shown at Table 1.

**Table 1.** The necessity of pre-job training courses (n=760)

| Necessity | Number of student | Proportion (%) |
| --- | --- | --- |
| Very necessary | 583 | 76.7 |
| Necessary | 173 | 22.8 |
| Not necessary | 4 | 0.5 |

### 4.2 Demand of pre-job training content

In order to understand the students’ needs for the content of pre-job training, we developed a multiple-choice questionnaire, the multiple-choice questionnaire main include 8 aspects (strengthening basic nursing skills, nursing safety and occupational protection, strengthening basic nursing theory, nurse-patient communication skills, introduction of basic information of practice hospital, regulations of nursing work, Nursing laws and regulations, Writing of nursing documents). The results showed that the most frequent choice was strengthening basic nursing skills (736 times, accounts for 96.8% in total 760 students), and then nursing safety and occupational protection (723 times, accounts for 95.1% in total 760 students), strengthening basic nursing theory (706 times, accounts for 92.9% in total 760 students), nurse-patient communication skills (670 times, accounts for 88.2% in total 760 students), introduction of basic information of practice hospital (658 times, accounts for 86.6% in total 760 students), regulations of nursing work (632 times, accounts for 83.2% in total 760 students), nursing laws and regulations (597 times, accounts for 78.6% in total 760 students), writing of nursing documents (547 times, accounts for 72.0% in total 760 students)(Table 2).

**Table 2.** Demand of pre-job training content (n=760)

| Pre-job training content | Frequency | Proportion in 760 students (%) |
| --- | --- | --- |
| Strengthening basic nursing skills | 736 | 96.8 |
| Nursing safety and occupational protection | 723 | 95.1 |
| Strengthening basic nursing theory | 706 | 92.9 |
| Nurse-patient communication skills | 670 | 88.2 |
| Introduction of basic information of practice hospital | 658 | 86.6 |
| Regulations of nursing work | 632 | 83.2 |
| Nursing laws and regulations | 597 | 78.6 |
| Writing of nursing documents | 547 | 72.0 |

### 4.3 Demand of nursing skills enhancement program

In order to understand the students’ needs for demand of nursing skills enhancement program, a multiple-choice questionnaire including closed intravenous infusion, Intradermal, subcutaneous and intramuscular injection, cardiopulmonary resuscitation, indwelling catheterization, closed venous blood transfusion, ECG monitoring, venous blood collection, nasogastric feeding, determination of vital signs, oxygen inhalation, aerosol inhalation, blood glucose measurement, oral care, seven-step hand-washing method was designed. The statistical results showed that the most frequent choice was closed intravenous infusion (686 times, accounts for 90.3% in total 760 students), and then intramuscular injection, cardiopulmonary resuscitation (667 times, accounts for 87.8% in total 760 students), cardiopulmonary resuscitation (622 times, accounts for 87.8% in total 760 students)(Table 3).

**Table 3.** Demand of nursing skills enhancement program (n=760)

| Nursing skills enhancement program | Frequency | Proportion in 760 students (%) |
| --- | --- | --- |
| Closed intravenous infusion | 686 | 90.3 |
| Intradermal, subcutaneous and intramuscular injection | 667 | 87.8 |
| Cardiopulmonary resuscitation | 622 | 81.8 |
| Indwelling catheterization | 589 | 77.5 |
| Closed venous blood transfusion | 586 | 77.1 |
| ECG monitoring | 580 | 76.3 |
| Venous blood collection | 546 | 71.8 |
| Nasogastric feeding | 536 | 70.5 |
| Determination of vital signs | 483 | 63.6 |
| Oxygen inhalation | 406 | 53.4 |
| Aerosol inhalation | 404 | 53.2 |
| Blood glucose measurement | 381 | 50.1 |
| Oral care | 368 | 48.4 |
| Seven-step hand-washing method | 240 | 31.6 |

### 4.4 Stress sources of pre-internship

A multiple-choice questionnaire including lack of confidence in theory and skills, distrust of patients, the strangeness of the environment, examination results and competition, clinical nursing teachers’ attitude towards themselves were designed for understanding the stress sources of pre-internship in 760 nursing students. the most frequent choice was lack of confidence in theory and skills, accounts for 61.1% in total 760 students, and then distrust of patients (accounts for 10.4% in total 760 students), the strangeness of the environment (accounts for 10.1% in total 760 students), those data showed at table 4.

**Table 4.** Stress sources of pre-internship

| Stress sources | Frequency | Proportion in 760 students (%) |
| --- | --- | --- |
| Lack of confidence in theory<br>and skills | 464 | 61.1 |
| Distrust of patients | 79 | 10.4 |
| The strangeness of the<br>environment | 77 | 10.1 |
| Examination results and<br>competition | 77 | 10.1 |
| Clinical nursing teachers' attitude<br>towards themselves | 63 | 8.3 |

## 5. Discussion

Pre-job training is an very important and essential content for nursing students in the early stage of clinical practice for nursing interns who just leave the classroom, get rid of the boring and tedious teaching of textbook knowledge, and are about to set foot on the clinical position in the hospital^[2]^. The content of pre-job training varies in different colleges and hospitals. According to the current pre-job training content for nursing students in major vocational colleges and the requirements of 2016 training program for new nurses issued by the National Health and Family Planning Commission(NHFPC), the pre-job training program for nursing students mainly includes three aspects: basic theoretical knowledge training, 26 items of common clinical nursing operation technology training, professional theory and practice ability training. Through the feedback of interns, practice hospitals and expert consultation, we developed a questionnaire survey on the pre-job training needs of nursing students in our school. Through the data analysis of the pre-job training needs of nursing students in our school, we could find the deficiencies of pre-job training and teaching in our school in time, and then combined with the actual situation of our school and the needs of practice hospitals, and modified the existing pre-job training system according to the survey results, to provide support for better clinical work of nursing students.

Pre-job training enables nursing students to master the basic theory, knowledge and skills of clinical nursing; cultivate good professional ethics, communication ability, emergency handling ability, professional care, condition observation, assistance treatment, psychological nursing, health education, rehabilitation guidance and other nursing service abilities required for implementing the responsibility system of holistic nursing; enhance humanistic care and sense of responsibility, can provide nursing services for patients independently and normatively^[3]^. The quality of clinical practice of nursing students is increasingly valued by schools and employers after graduation, and even has a lifelong impact on the nursing work of nursing students after graduation^[4]^. Scientific and standardized pre-job training can quickly improve the deficiencies of students before practice, strengthen the nursing theory and skills operation ability, ensure the safety of students’ practice, and improve the quality o f nursing teaching and social satisfaction.

In this convey, there were 583 students thinking that pre-job training was very necessary, accounts for 76.7% of the total number of students surveyed; According to the multi-choice questionnaire results, the most frequent choice was strengthening basic nursing skills (736 times, accounts for 96.8% in total 760 students), and then nursing safety and occupational protection (723 times, accounts for 95.1% in total 760 students), strengthening basic nursing theory (706 times, accounts for 92.9% in total 760 students), those data showed that strengthening basic nursing skills, nursing safety and occupational protection and strengthening basic nursing theory is of the top priority. Schools should fully consider the needs of nursing students in school, pre-job training content as close to clinical nursing as possible; at the same time, the pre-job training course should be integrated into the core system of nursing specialty, the relevant guidance of laws and regulations, strengthen students’ legal awareness.

For designing the questionnaire content of the nursing skills enhancement program, this study combined with the feedback of interns, nursing expert interviews, 2016 new nursing students training program, prepared 14 common clinical nursing operations. the most frequent choice was closed intravenous infusion (686 times, accounts for 90.3% in total 760 students), and then intramuscular injection, cardiopulmonary resuscitation (667 times, accounts for 87.8% in total 760 students), cardiopulmonary resuscitation (622 times, accounts for 87.8% in total 760 students). According to the results, the school should focus on the operation items with the most needs of students for intensive training and guidance. The rest of the operations can be practiced by video learning+open training room practice+teacher guidance. In the skill intensive training, teachers in higher vocational colleges should be close to the actual clinical requirements, analyze and guide the difficulties and key points of various operations, and emphasize the importance of hand washing, aseptic technology and medical waste classification for the prevention of nosocomial infection, so as to realize effective pre-job training and intensive training.

In the survey, 61.1% of the students’ pre-internship pressure mainly comes from their own theory and skills are not confident, which fully demonstrated that the importance and necessity of pre-job training. The establishment of scientific and standardized pre-job training courses and the close communication between the school and the internship cooperation hospital can meet the physical and mental needs of students before clinical practice, help students complete the role transformation, and have the confidence and ability to enter the clinical practice.

Pre-job training is a summary of two or three years’ theory and skill learning of nursing students, strict pre-job training management is responsible for nursing students and patients. In this paper, we investigated the necessity of pre-job training for nursing students, the demand of pre-job training content, the demand of nursing skill enhancement program and the stress source of pre-practice in our school, those data may have practical guiding significance and provide scientific basis for the formulation of pre-job training content and training program in our school.

## Data Availability

The data used to support the findings of this study are available from the corresponding author upon request.

## 7. Acknowledgements

None.

## References

[1] Deng Shengwei. Exploration and Research on standardized pre-job training mode for new nurses [J]. Contemporary nurses (In Chinese), 2019, 26 (08): 170–171

[2] Shao Yanrong, Qian Hongying. Investigation report on pre-job training demand of nursing interns [J]. Massage and rehabilitation medicine(In Chinese), 2018, 9 (21): 75–77

[3] Chinese National Health Office (2016) No. 2, training program for new nurses in 2016.

[4] Zhang Yanjie, Chen Xiuyun, Zhang Yiwen, Ni Feng, Liu pengmei. Research progress on Countermeasures to improve the quality of nursing students’ practice [J]. Nursing practice and research (In Chinese), 2015, 12 (04): 21–23

